# Patterns of deprescribing after an emergency department visit due to adverse drug events among concomitant users of antithrombotics and other medications

**DOI:** 10.64898/2026.07.20.26358502

**Authors:** Emily Chi, Ahmed Soliman, Katherine M. Hunold, Chien-Wei Chiang, Kathleen T. Unroe, Regina N. Nechi, Jeffrey M. Caterino, Lang Li, Pengyue Zhang, Macarius Donneyong

## Abstract

**Objectives:** This study aimed to examine patterns of deprescribing practices that are implemented after an emergency department (ED) visit due to antithrombotic-induced gastrointestinal (GI) bleeds among patients who concomitantly use antithrombotic and other medications.

**Methods:** A retrospective cohort study design was used to identify a cohort of patients who visited an ED from the MarketScan® claims database (2016 – 2023). Concomitant use of antithrombotics and other medications was assessed in the 30 days prior to the ED date (index date). Those who presented with GI bleeds on their ED visit were classified as exposed vs those without GI bleeds (unexposed).

Antithrombotic deprescribing – defined as discontinuation (≥45 days gap between refills), switching, or dose reduction – was assessed from the index date through to end of data. Inverse probability of treatment (IPT) – weighted logistic regression models were used to compare the odds of deprescribing between the exposed vs. unexposed groups.

**Results:** Of the 375,510 total concomitant users of antithrombotics and other agents, 9,145 had a GI bleed vs. not (366,365). The odds of antithrombotic deprescribing was significantly higher among the exposed vs unexposed, (odds ratio [OR], 1.39; 95% confidence interval [CI], 1.33, 1.46). There was no significant difference in the discontinuation of concomitant medications overall. However, among patients who continued antithrombotic use after the ED visit, the discontinuation of concomitant medications was relatively higher among the exposed (OR, 1.11; 95% CI, 1.05, 1.16).

**Conclusions:** Antithrombotic agents were more likely to be deprescribed after an ED visit among patients who concomitantly used antithrombotics and other medications.

**Study Highlights:** *WHAT IS KNOWN:*

- The use of antithrombotic agents alone is associated with increased risks of gastrointestinal (GI) bleeding, and this may be exacerbated by the concurrent use of other medications.
- Deprescribing (i.e. discontinued, switched, or dose reduction) of antithrombotics after GI bleeding events is commonly practiced.

*WHAT IS NEW HERE:*

- This was the first study to comprehensively examine deprescribing practices of antithrombotic and concomitant medications following ED visits among patients with versus without GI bleed.
- Among patients who concomitantly use antithrombotic and other agents, clinicians were more likely to deprescribe the non-antithrombotic concomitant medications after an ED visit due to a GI bleed.
- The most commonly co-prescribed medications with antithrombotics were antihypertensives and statins, the discontinuation of at least one medication in the three-drug combinations was more common among exposed than unexposed.

## Introduction

Antithrombotics, an umbrella term that includes both anticoagulants and antiplatelet agents, are commonly used for treatment and prevention of myocardial infarction, stroke, and venous thromboembolism.^1–3^ However, bleeding, especially in the gastrointestinal (GI) tract, is a major adverse event of antithrombotic treatment and is associated with nearly 80% increase in emergency department (ED) visits, and up to 10% increased risk of death.^4,5^

When a patient presents to the ED with an adverse drug event (ADE), reversal or temporary interruption of the potentially damaging drug is a recommended clinical practice.^6–8^ This is known as deprescribing, defined as “the process of clinician-supervised medication withdrawal” through discontinuation, dose change, particularly dose reduction, and medication switching for a safer alternative.^9–11^ Per the American College of Gastroenterology (ACG) guidelines, some level of deprescribing of antithrombotic agents is encouraged after an acute GI bleeding event.^7^ Some studies have reported that short-term discontinuation of antithrombotic agents after an acute GI bleeding event is safe and does not increase the risk of stroke or systemic embolism.^12^

Despite the known risks of GI bleeding among patients concomitantly exposed to antithrombotic agents and other medications, polypharmacy is common among users of antithrombotics. Approximately 66% of antithrombotic users concurrently use five or more medications,^13^ increasing the risk of DDIs and complicating clinical management.^14,15^

Current research on deprescribing has been mostly focused on methods to implement deprescribing protocols in different settings, such as the ED,^16^ or in specific groups of people, such as older adults with cardiovascular disease,^17^ implementation barriers,^18^ in a particular class of medication,^19^ or in a particular healthcare system.^20^ Another major knowledge gap in the published literature is the lack of data on how frequently clinicians deprescribe antithrombotic agents and/or concomitant medications after a GI bleeding event. To address this knowledge gap, we sought to examine the deprescribing practices that are often implemented after an ED visit due to a GI bleeding event using real world data. Specifically, we sought to quantify the likelihood of the discontinuation of antithrombotics (with or without concomitant medications), switching between antithrombotic agents, or a reduction of the dose of an antithrombotic agent.

## Methods

### Data source

We used the Merative® MarketScan® health insurance claims data from January 2016 to December 2023. The MarketScan® dataset contained individual-level healthcare claims, including inpatient, outpatient, and pharmacy claims. The use of this data was approved by The Ohio State University IRB.

### Study design and cohort selection

A retrospective cohort study design was used to examine the deprescribing of antithrombotic and concomitant medications among patients in ED settings (Figure 1). Exposed group was defined as patients with at least one ED visit that had a record of GI bleeding in the study period; unexposed group was defined as patients who had no ED visits record of GI bleeding. The index date for a patient was defined as their first ED visit with a GI bleed for the exposed group, and the first ED visit without a GI bleed for the unexposed group. ED visits were identified using Healthcare Common Procedure Coding System (HCPCS) codes (99281–99285), revenue codes, and place of service (i.e. place of service = ED). GI bleeding was identified using International Classification of Disease Ninth and Tenth Revisions (ICD-9, ICD-10) codes. We identified patients who used an antithrombotic, defined as medications under B01A-Antithrombotic agents in the Anatomical therapeutic chemical (ATC) classification (Supplement Table 1) within 30 days before ED visit. These patients were also required to be using at least one other type of medication that overlapped with the antithrombotic within 30 days before ED visits and to have continuous enrollment in health insurance plan 180 days before ED (Figure 2).

**Figure 1.**
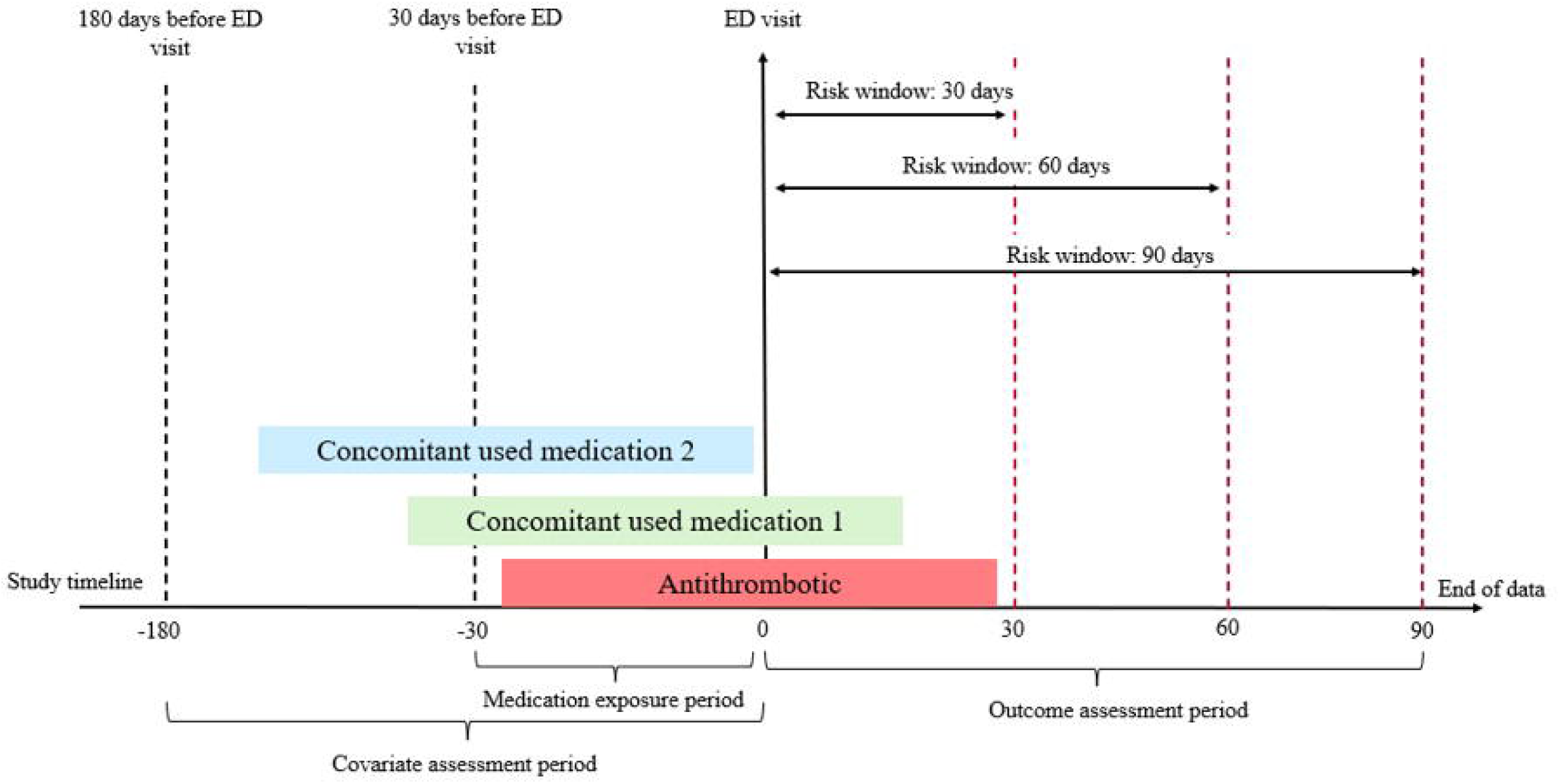
Study framework and operational definitions for deprescribing. To examine the deprescribing of antithrombotics and concomitant medications in emergency department (ED) setting, the first ED visit was defined as index date. Covariates were assessed within 180 days before the ED visit. The medication exposure period included antithrombotic therapy and concomitant medications used within 30 days before the ED visit. Following the ED visit, outcomes were assessed during post-ED risk windows of 30, 60, 90 days, and through the end of data availability. Abbreviations: emergency department = ED.

**Figure 2.**
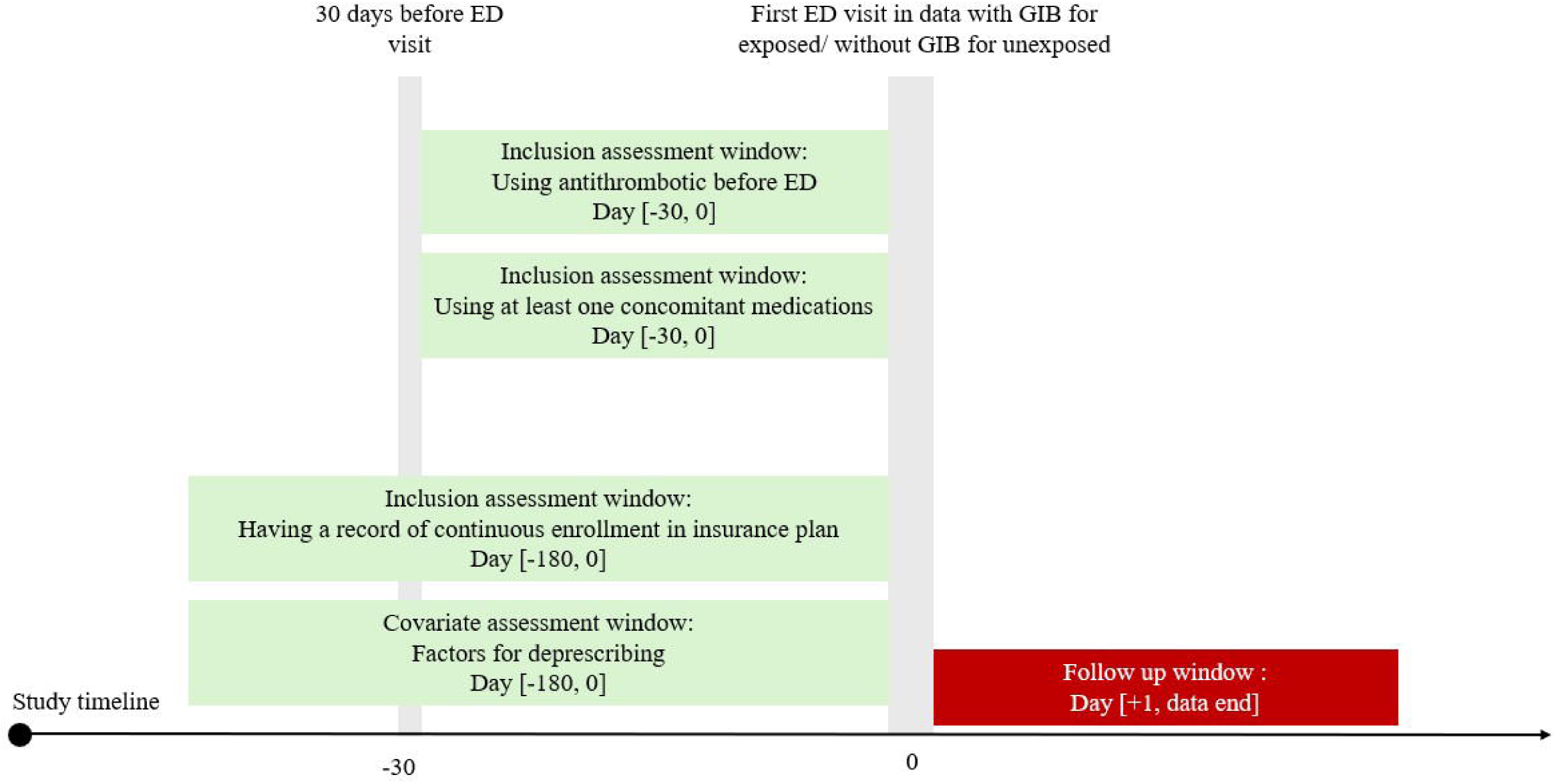
Descriptions of cohorts of patients using antithrombotic and concomitant medications presenting to the emergency departments. Inclusion criteria were patients who received antithrombotics and at least one other type of medication within 30 days before emergency department (ED) and had at least 180 days of continuous enrollment in health insurance plan before the ED visit. Patients with gastrointestinal bleed (GIB) during the first ED visit in the dataset were classified as exposed and those without GIB were classified as unexposed. Abbreviations: emergency department = ED; gastrointestinal bleed = GIB.

### Outcome measurement

Patients were followed from the index date until the end of data availability for deprescribing events: (1) discontinuation, (2) switching, or (3) dose reduction. Medication (antithrombotic or concomitant medication) discontinuation was deemed to have occurred if there was an observed gap of ≥ 45 days between the last date of the day’s supply of a drug and the next refill date of the same drug (if refilled), or end of data availability, whichever comes first.^21,22^ Switching (applied to only antithrombotics) was defined as a change from the use of one antithrombotic in the baseline period to a different one. Dose reduction was defined as a decrease in the strength of antithrombotic prescribed before ED visits. Additionally, different risk windows were defined (30, 60, or 90 days after the ED visit) to assess the timing of deprescribing events. Deprescribing events were evaluated within each risk window based on whether the deprescribing date fell within the specified time frame following the ED visit.

### Covariates

All baseline covariates were measured within 180 days before ED visit. We identified demographic variables, comorbidities, and overall health status, summarized using Charlson comorbidity index, CHA_2_DS_2_-VASC, and HAS-BLED scores. Healthcare utilization was assessed by the number of outpatient visits and encounters, hospitalizations, length of stay, and number of concomitant medications used within 30 days before the ED visits. Details were provided in Supplemental Table 2.

## Statistical analysis

Given the potential for baseline difference between patients with and without GI bleeds, we used inverse probability of treatment weighting (IPTW) to balance covariates between the exposed and unexposed groups.^23^ We calculated each patient’s propensity scores as the predicted probability of being exposed to GI bleeding, conditional on all baseline covariates. Standardized mean differences (SMD) were used to assess balance of covariates before and after IPTW.

Baseline characteristics and descriptive outcomes were summarized using means with standard deviation for continuous variables, and frequencies (counts with percentages) for categorical variables. In the main analysis, we described the deprescribing events among exposed vs. unexposed using weighted count and percentage by following ED visits until the end of data availability. We also described these patterns using different risk windows (30, 60, or 90 days after the ED) to better understand whether deprescribing events were related to the ED visits. To further understand changes in the frequency of specific types of antithrombotic and concomitant medication use and deprescribing before and after the ED visit, frequencies were used to describe medication use and deprescribing events. Bar charts compared antithrombotic use before ED visits and deprescribing after ED visits between groups. The Sankey plot was used to illustrate the top 10 frequently observed three-drug combinations (one antithrombotic and two concomitant medications) in our analytic cohort, and medication discontinuation trajectories within drug combinations among exposed and unexposed groups.

To compare the prevalence of antithrombotic deprescribing events among exposed vs unexposed, we applied logistic regression models to calculate the odds ratio (OR) and 95% confidence intervals (CI) for antithrombotic deprescribing. These models were weighed by inverse probability of treatment (IPT) to balance distribution of the measured covariates between two groups. To evaluate the individual antithrombotic deprescribing events, separate IPT-weighted logistic regression models were conducted for each type of deprescribing event (discontinue, switching, and dose reduction). Similarly, we used IPT-weighted logistic regression to calculate the ORs and 95% CIs for concomitant medication discontinuation in the exposed vs unexposed groups among patients by antithrombotic deprescribing and antithrombotic discontinuation status. All the ORs and 95% CIs from IPT-weighted logistic regression were presented in forest plots. Statistical analyses were conducted using SAS, version 9.4. Two-sided p-values at an alpha of 0.05 were used to determine statistical significance.

## Results

A total of 375,510 patients with ED visits were included, including 9,145 patients with GI bleeds (exposed) and 366,365 patients without GI bleeds (unexposed). After applying IPTW, the weighted sample included 8,691 exposed and 366,249 unexposed individuals, resulting in a balanced distribution of measured covariates between the exposed vs unexposed (Table 1). The mean age was 68.6 years in the exposed group and 65.8 years in the unexposed group.

**Table 1.**
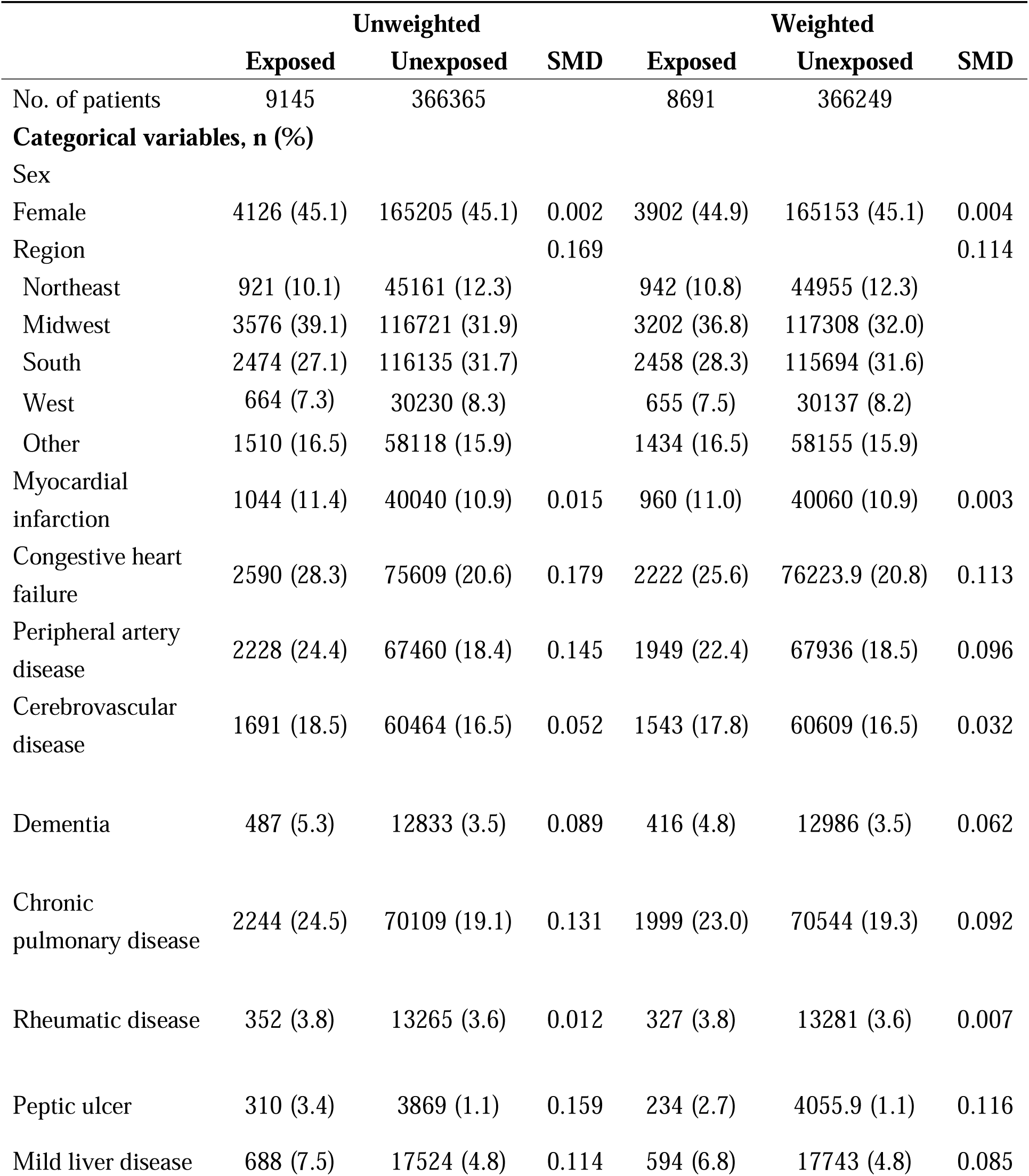
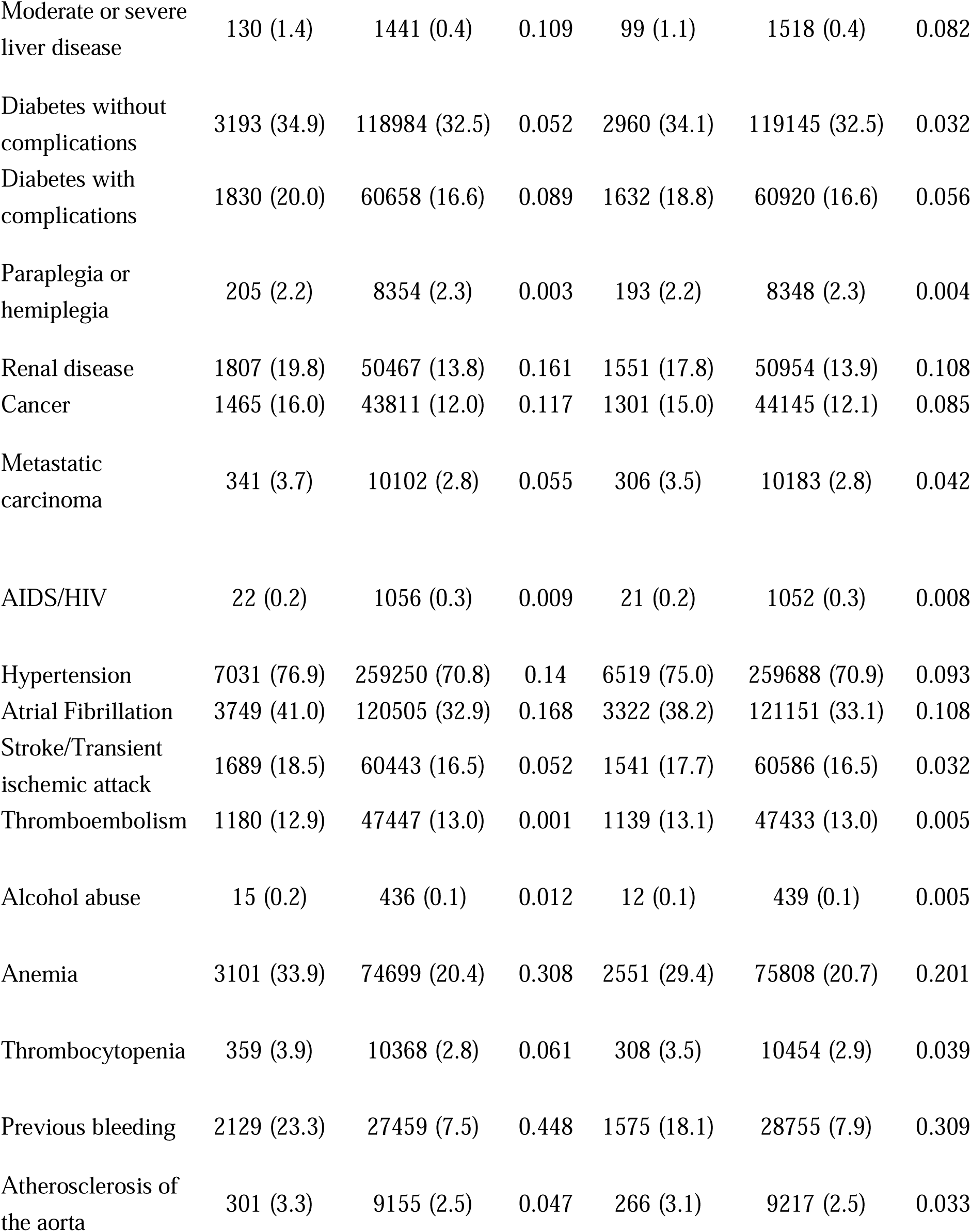
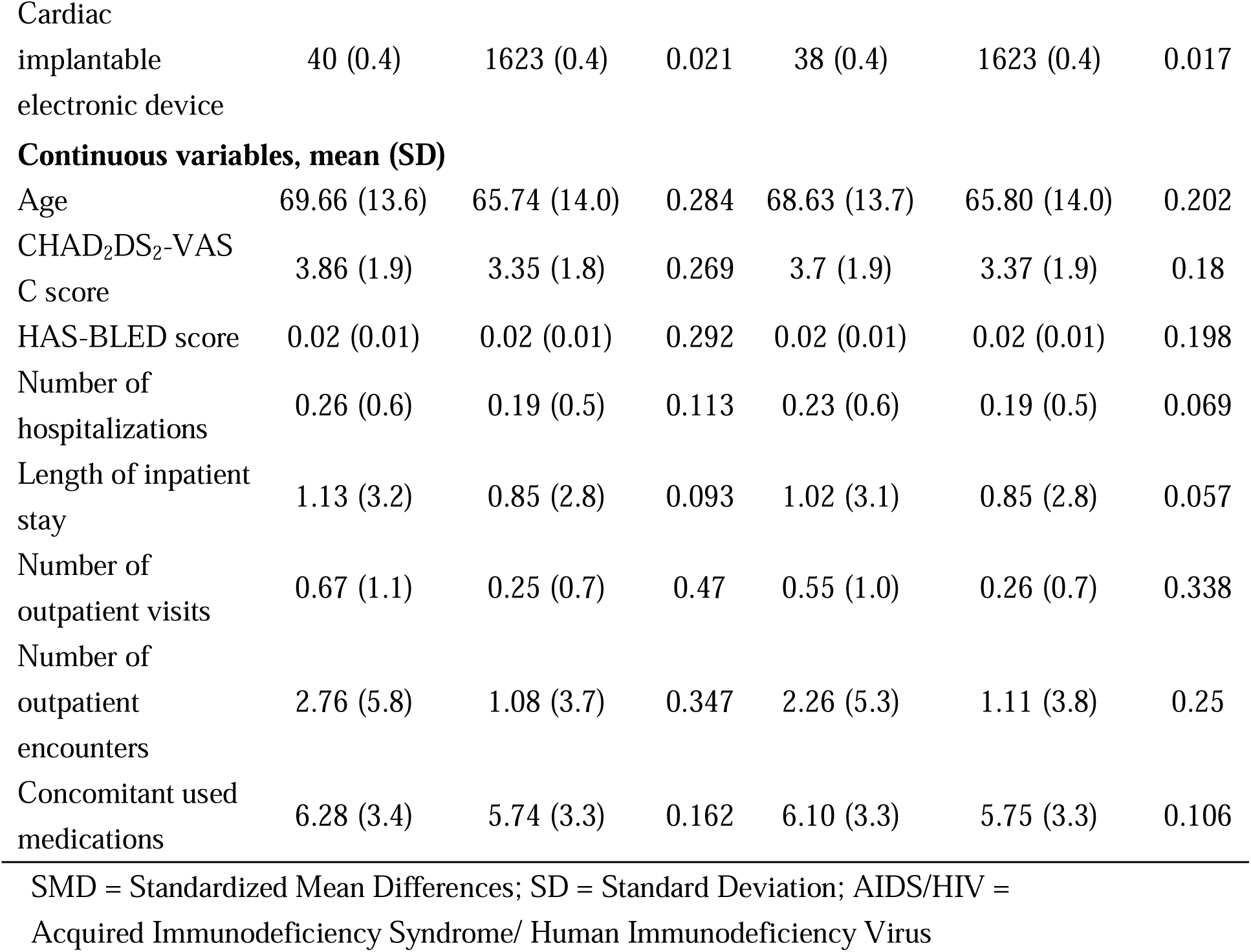
Baseline characteristics of patients before and after inverse probability of treatment weighting.

The frequency and patterns of deprescribing events are shown in Figure 3. Overall, 33.0% of exposed vs. 26.2% of unexposed had at least one antithrombotic deprescribing event after an ED visit: 18.8% discontinued (23.6%, vs 18.7%), 5.7% switched (7.1% vs 5.6%), 1.9% had a dose reduction (2.4% vs 1.9%). The exposed group had higher percentage of antithrombotic deprescribing than unexposed within 30 days (13.9% vs 11.5%), 60 days (19.5% vs 14.9%), and 90 days (23.5% vs 17.3%), with deprescribing events increasing across longer risk windows.

**Figure 3.**
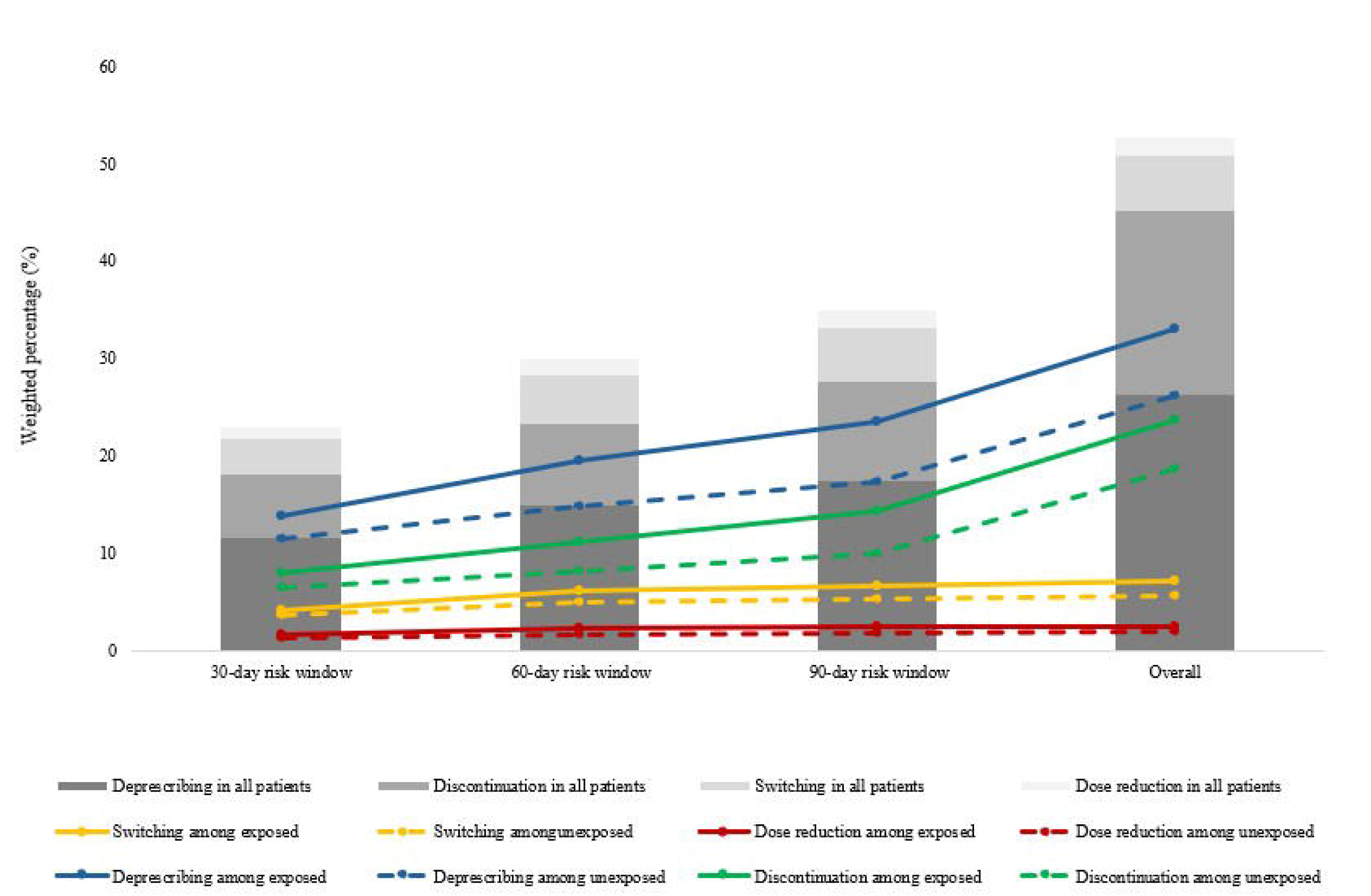
Distribution of antithrombotic deprescribing and different components of deprescribing among exposed and unexposed across different risk windows. This figure illustrates the percentage of antithrombotic deprescribing and components (discontinuation, switching, and dose reduction) across 30-day, 60-day, and 90-day risk windows following the index emergency department visits, and the overall study period. Across all windows, the exposed group consistently showed higher percentages of deprescribing compared to the unexposed group.

Patients with GI bleeds had 39% higher odds of antithrombotic deprescribing (OR, 1.39; 95% CI, 1.33, 1.46; *P* <.001) compared with those without GI bleeds (Figure 4A). Examining the unique deprescribing events, the odds of discontinuation (OR, 1.38; 95% CI, 1.27, 1.50; *P* <.001), switching (OR, 1.39; 95% CI, 1.32, 1.46; *P* <.001), and dose reduction (OR, 1.42; 95% CI, 1.23, 1.63; *P* <.001) were significantly higher among patients with GI bleeds than those without GI bleeds (Figure 4A).

**Figure 4.**
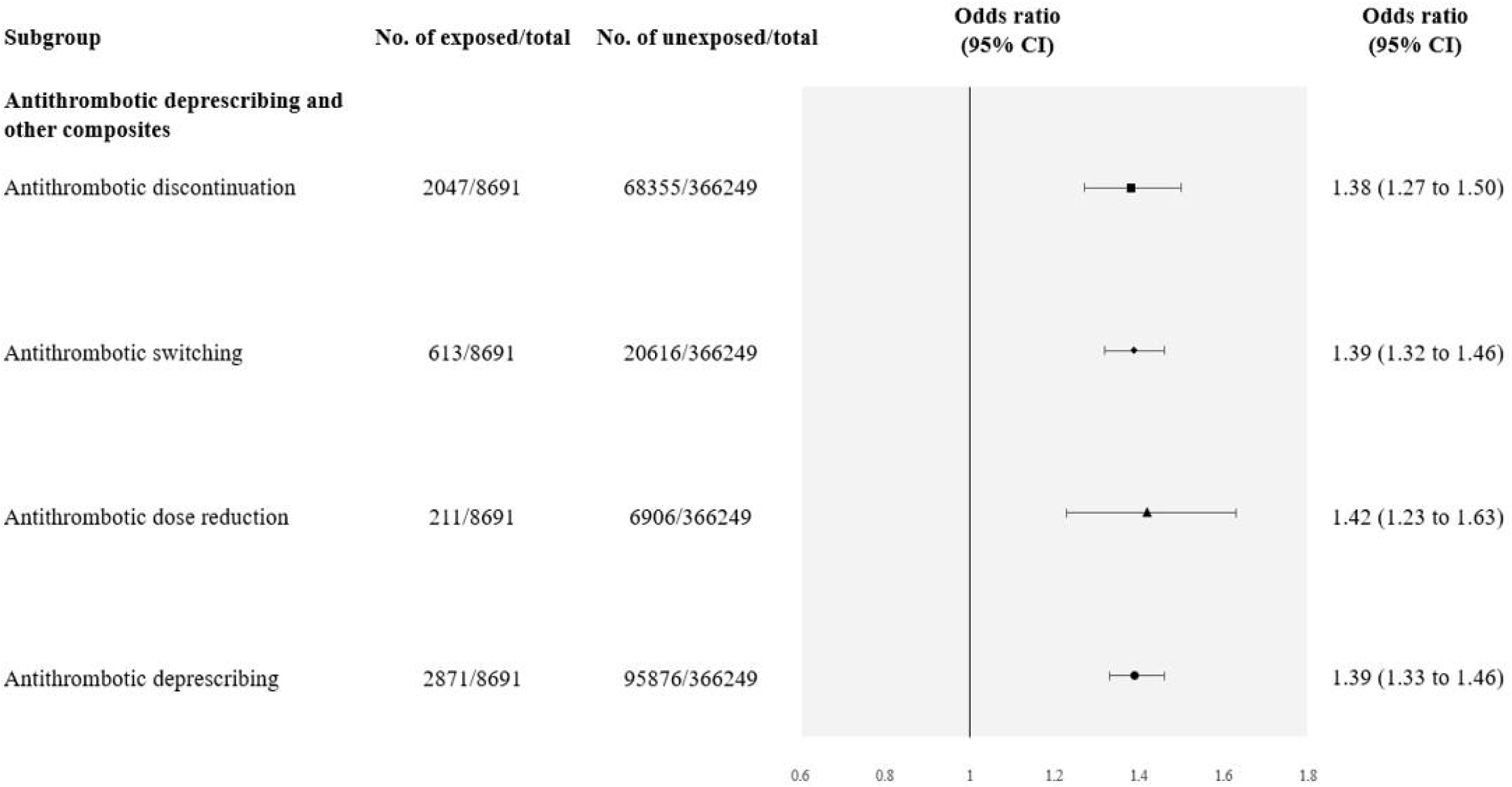
(A) Odds ratios for antithrombotic deprescribing and different components of deprescribing among exposed vs unexposed; (B) Odds ratios for concomitant medication discontinuation stratified by whether patients had antithrombotic deprescribing and discontinuation among exposed vs unexposed. The forest plots illustrate the odds of antithrombotic deprescribing and concomitant medication discontinuation. Compared to unexposed, the exposed had higher odds of antithrombotic deprescribing and other components of deprescribing. There was no significant difference between the exposed and unexposed among patients with antithrombotic discontinuation. However, among patients continuing antithrombotic, exposed had higher odds of concomitant medication discontinuation than unexposed. Abbreviations: 95% Confidence Interval = 95% CI.

In Figure 4B, we reported the ORs for the concomitant medication discontinuation by the subgroup of patients whose antithrombotics were deprescribed vs not. Among patients with antithrombotic deprescribing, concomitant medication discontinuation did not significantly differ between the exposed vs unexposed (61.2% vs 61.0%), (OR, 1.01; 95% CI, 0.94, 1.09; *P*=.82). However, the odds of concomitant medication discontinuation was higher among the exposed (62.4%) vs unexposed (63.6%) among patients without antithrombotic deprescribing (OR, 1.09; 95% CI, 1.04, 1.15; *P* <.001).

Figure 4B showed the ORs for the concomitant medication discontinuation by whether patients antithrombotics were discontinued. Among patients with antithrombotic discontinuation, concomitant medication discontinuation did not significantly differ between exposed vs unexposed (62.4% vs 63.6%), (OR, 0.95; 95% CI, 0.89, 1.04; *P*=.26). In contrast, among patients without antithrombotic discontinuation, the odds of concomitant medication discontinuation was higher in exposed (54.5%) than unexposed (52.1%), (OR, 1.11; 95% CI, 1.05, 1.16; *P* <.001) (Figure 4B). When we restricted to 30-, 60-, and 90-day risk, the exposed consistently had a higher percentage of concomitant medication discontinuation than the unexposed group, with discontinuation increasing across longer risk windows (Supplement Table 3).

Figure 5 showed the change in antithrombotic agents before and after ED visit in exposed vs unexposed. Among identified antithrombotic in our study, the top four most frequently prescribed antithrombotics before ED visits were similar between the exposed vs unexposed: clopidogrel (29.2% vs 30.6%), apixaban (26.2% vs 26.3%), rivaroxaban (17.4% vs 14.5%), and warfarin (15.4% vs 13.9%). After ED visit, the exposed group had higher percentage of deprescribing on clopidogrel (9.1% vs 6.8%), apixaban (7.3% vs 5.3%), rivaroxaban (5.0% vs 3.5%), and warfarin (6.5% vs 4.7%) than unexposed.

**Figure 5.**
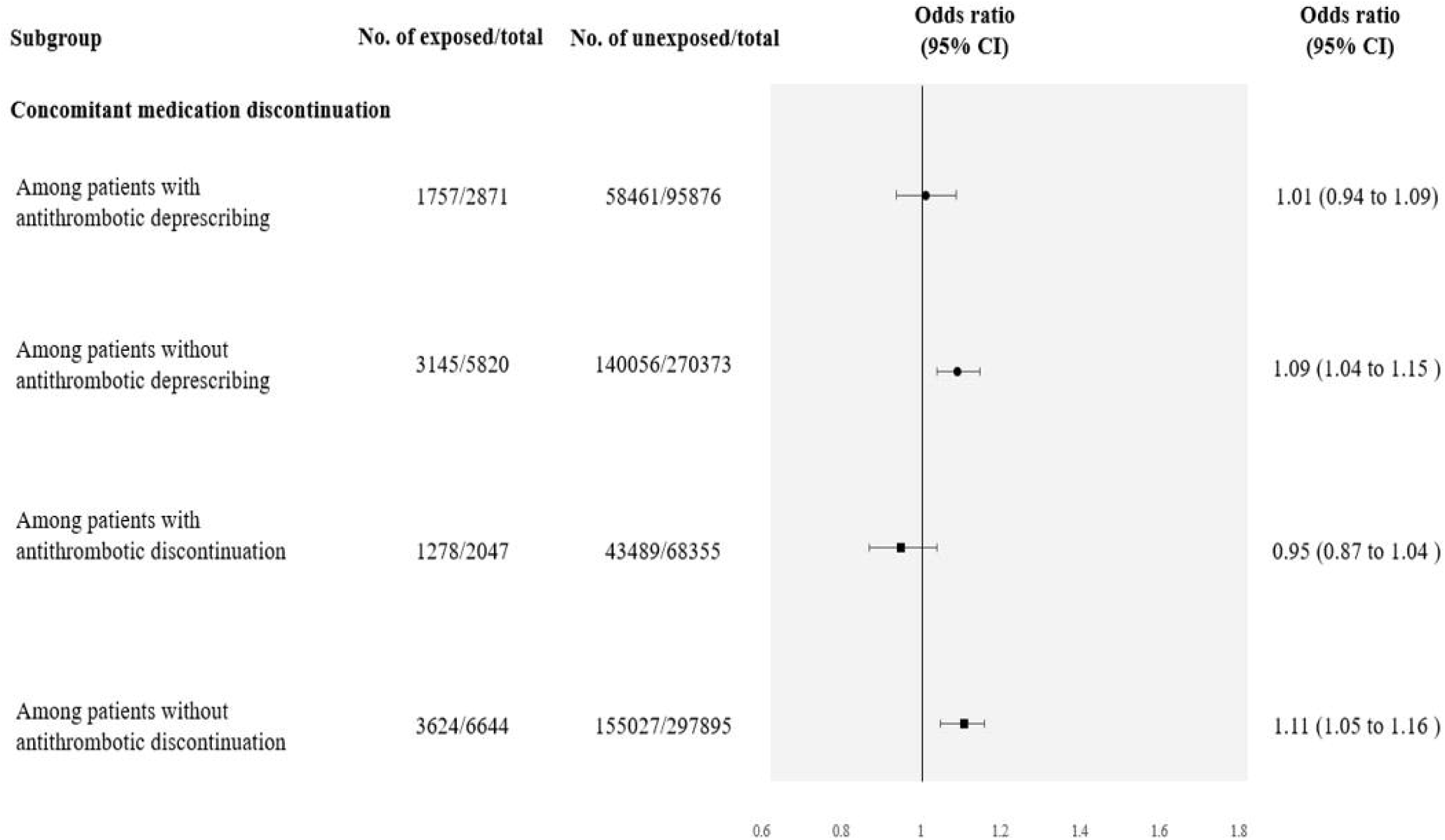
Change in distribution of antithrombotic use before ED visit and post-ED deprescribing among exposed vs unexposed. Bar charts were generated to compare the distribution of antithrombotic use before the ED visit and deprescribing events after the ED visit between the exposed and unexposed groups. The top four most frequently prescribed antithrombotics (clopidogrel, apixaban, rivaroxaban, and warfarin) before ED visits were similar between the exposed vs unexposed. After ED visit, exposed group had higher percentage of deprescribing than unexposed among these four antithrombotic agents. Abbreviations: ED = emergency department.

Supplement Table 4-5 showed the top 20 most frequently used concomitant medications and discontinuation pattern. Among exposed vs unexposed group, metoprolol (35.1% vs 35.0%) and atorvastatin (35.0% vs 34.7%) were the most frequently used concomitant medication before ED visits. However, furosemide was more commonly used among exposed (17.0%) than unexposed (13.9%). After the ED visit, discontinuation of metoprolol and atorvastatin were similar between exposed vs. unexposed: metoprolol (7.6% vs 7.6%) and atorvastatin (3.5% vs 3.7%). In contrast, furosemide discontinuation was higher among exposed (3.9%) than unexposed (3.1%).

Figure 6 showed the top 10 frequently used three-drug combination and discontinuation among exposed vs unexposed. The most frequent combinations in both groups before ED visit generally consisted of an antithrombotic, antihypertensive, and statin. Clopidogrel + metoprolol + atorvastatin was the most common combination in both groups (5.5% exposed vs 6.0% unexposed). Following the ED visit, exposed had a higher percentage of at least one medication discontinuation within the combination. Among those using clopidogrel + metoprolol + atorvastatin, 38.6% of exposed discontinued at least one medication compared with 33.5% of unexposed. Discontinuation of clopidogrel was also higher among exposed (22.2%) than unexposed (14.5%). Similarly, among patients using apixaban + metoprolol + atorvastatin, 34.1% of exposed discontinued at least one medication, compared with lower percentage of discontinuation observed in comparable combinations among unexposed patients (31.9%). Across other frequent combinations, exposed patients consistently demonstrated higher percentages of discontinuation of at least one medication (exposed: 32–46% vs. unexposed: 27–37%) and higher percentages of antithrombotic discontinuation (exposed: 20–28% vs. unexposed: 14–16%) (Supplement Tables 6–7).

**Figure 6.**
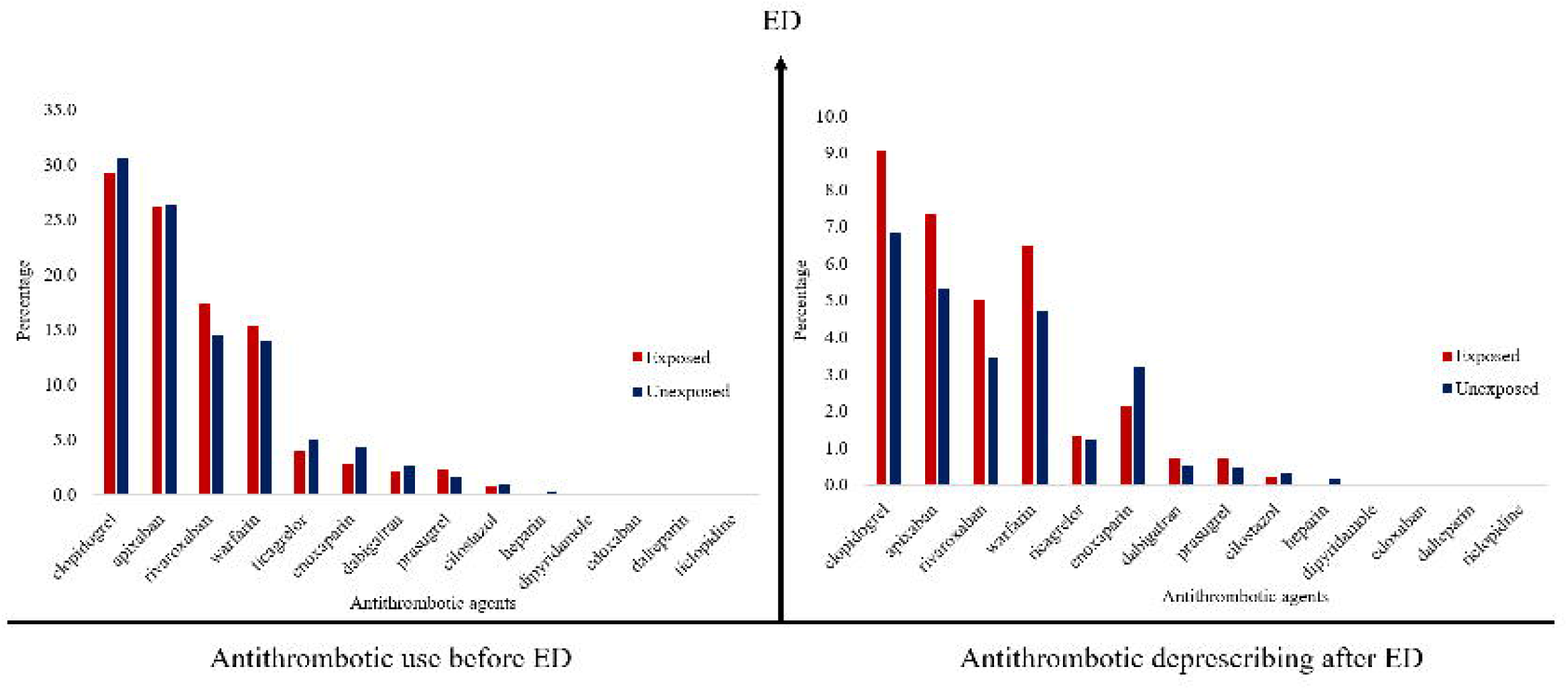
Top 10 frequently used three-drug combination and discontinuation among exposed and unexposed groups. The Sankey diagram illustrates the medication discontinuation trajectories for the top 10 most frequent three-drug combinations, comparing between exposed and unexposed. The combinations frequently consist of an antithrombotics paired with statins and antihypertensives. Compared to the unexposed group, the exposed group exhibited a higher percentage of both antithrombotics and concomitant medication discontinuation.

## Discussion

In this large commercially insured cohort of patients who concomitantly used antithrombotics and other medications (most commonly antihypertensives or statins) before an ED, antithrombotic agents were more likely to be deprescribed among those with GI bleed compared to those without. We observed that patients were more likely to discontinue concomitant medication if antithrombotics were maintained after an ED visit due to a GI bleed.

Previous studies reported antithrombotic discontinuation rates of 41-58% following GI bleeding.^24–26^ In contrast, we observed a lower discontinuation rate (23.6%) among patients with GI bleeds. Those studies defined discontinuation based on the resumption of anticoagulant therapy after GI bleed, classifying patients who did not restart therapy during follow-up period as discontinuing anticoagulants.^24–26^ In contrast, we defined discontinuation using gaps between refills following ED visits. Those studies have primarily focused on warfarin, and their average age range was 74.2-78.4. In our cohort, only 15.5% of patients with GI bleeds (average age: 68.6) received warfarin prior to ED visits, whereas 34.3% received DOACs, which may contribute to lower discontinuation rates.

No published studies examining the odds of antithrombotic deprescribing and concomitant medication discontinuation after ED visits. While antithrombotic deprescribing was more frequent among exposed (patients with GI bleeds) than unexposed (patients without GI bleeds), we found no significant difference in the odds of concomitant medications discontinuation between exposed and unexposed among those with antithrombotic discontinuation. In contrast, when antithrombotic therapy was continued, exposed were more likely to discontinue concomitant medications than unexposed.

Once antithrombotic therapy is discontinued, the primary driver of bleeding risk is removed and additional medication changes may offer limited benefit. However, among patients who continued antithrombotic therapy, concomitant medication adjustment may represent an alternative approach to reduce overall bleeding risk. Importantly, after a GI bleed, continuing or discontinuing antithrombotic medications may be appropriate depending on patient’s risk.^27^ Because this is a claims database study, we cannot determine clinicians’ decision-making; however, these patterns may reflect individualized balancing of bleeding and thrombotic risks.

Our study identified several concomitant medications that have been reported to interact with antithrombotic agents and increase bleeding risk. When examining potential drug-drug interactions (DDIs) between the top 20 concomitantly used medications and four most commonly used antithrombotic agents (warfarin, clopidogrel, rivaroxaban, and apixaban) before the ED visits in our study, we found that allopurinol may enhance anticoagulant effect of warfarin, increasing the risk of bleeding.^28,29^ In our study, allopurinol discontinuation was observed only among exposed, with 0.6% experiencing discontinuation. Diltiazem has been associated with 19% higher odds of GI bleeding in a meta-analysis when used with DOACs.^30^ Our study found that both exposed and unexposed had low discontinuation for diltiazem after ED visits (0.8% vs 0.6%). Other concomitant medications may potentiate the anticoagulant effect of warfarin such as levothyroxine sodium, rosuvastatin, and simvastatin. Concomitant use of such medications may require closer monitoring of the patient’s INR and adjusting the warfarin dose accordingly.^31–33^ In terms of drug combinations, one frequent combinations in the GI bleeding group included clopidogrel + pantoprazole + atorvastatin, has been associated with high risk of GI bleeding.^34^ Moreover, switching from pantoprazole to dexlansoprazole was found to be a clinically acceptable alternative with lower risk of GI bleeding.^35^

### Clinical implications

Overall, patients were more likely to have an antithrombotic agent deprescribed after an ED visit for GI bleed. This data reassures that clinicians are following the ACG guidelines.^7^ Interestingly, when the antithrombotics agents were maintained after a GI bleed, it appears that clinicians often opted to discontinue the concomitant medications instead. This may reflect clinical trade-off between maintaining antithrombotics and discontinuing concomitant medications. Clinicians should weigh the balance between bleeding and thrombotic risk when making deprescribing, particularly in acute care setting.^36^ This is because all antithrombotic agents carry an increased risk of bleeding and are frequently linked to ED visits related to ADEs.^37^ GI bleeding is recognized as a common complaint in the ED, accounting for over 600,000 ED visits annually and contributing to 17% of ADEs in ED.^38,39^ Despite the frequency and clinical importance of these events, empirically established deprescribing protocols to guide clinicians remain limited.^40^

Prior research demonstrated that although bleeding and thrombotic events occur at similar frequencies, mortality after thrombotic events is nearly twice that after major bleeding,^41^ and 44% of patients at high bleeding risk may simultaneously remain at high thrombotic risk.^41^ This shows that deprescribing antithrombotic medications is a nuanced decision where the risk of bleeding must be weighed against the risk of thrombotic events. In this circumstance, clinicians may consider maintaining antithrombotic therapy for patients with high thrombotic risk despite a recent GI bleed, instead prioritizing the discontinuation of concomitant medications to mitigate the overall bleeding risk.^42^

Based on our data, we identified several concomitant medications and drug combinations that have previously been reported to interact with antithrombotics and increase GI bleeding risk. The concurrent use of multiple medications increases the risk of ADE due to potential DDIs. Beyond discontinuation of concomitant medications, our prior studies have suggested substituting high-risk drugs with lower-risk alternatives that maintain therapeutic efficacy.^35^ Together, these findings suggest that deprescribing following ED visit is balanced against patients bleeding and thrombosis risk.

### Strengths & limitations

Our study has several limitations. First, it was a retrospective study using claims data, so we could not capture medications obtained without insurance claim or administered in certain settings (e.g., inpatient medication administration). Moreover, claims data don’t reflect patients’ actual medication-taking behavior or physicians’ clinical reasoning behind deprescribing decisions. Despite these limitations, our study also has several strengths. First, we used a large-scale, real-world dataset, providing comprehensive and generalizable samples. Second, we examined multiple deprescribing outcomes (discontinuation, switching, and dose reduction) allowing better understanding of medication changes after ED. Lastly, we applied IPTW to mitigate confounding and improve the comparability between the exposed and unexposed groups.

## Conclusion

In conclusion, in a large population of patients concurrently using antithrombotic agents and other medications, presenting GI bleeding in the ED versus others without GI bleed had higher odds of antithrombotic deprescribing. Concomitant medications tended to be discontinued when antithrombotics were continued, suggesting clinical trade-off between bleeding and thrombotic risks.

## Conflict of interest

### Guarantor of the article

Macarius Donneyong is the guarantor of this article

### Specific author contributions

Emily Chi contributed to the conceptualization and design of the study, data analysis and interpretation, and manuscript drafting. Ahmed Soliman contributed to the conceptualization and design of the study and manuscript drafting. Chien-Wei Chiang contributed to the acquisition and analysis of the data. Katherine M. Hunold, Kathleen T. Unroe, Jeffrey M. Caterino, Lang Li, and Pengyue Zhang contributed to the development of the design of the study, data acquisition, and manuscript drafting. Regina Nechi contributed to manuscript review. Macarius Donneyong contributed to the conceptualization and design of the study, data analysis and interpretation, data acquisition, and manuscript drafting.

### Financial support

The study was supported by the NIA grant R01AG071018. Dr. Hunold was receiving support from NIA grant K76AG074941.

### Potential competing interests

The authors declare no conflict of interest.

## Data Availability

The data that support the findings of this study are available from Merative MarketScan under license and are not publicly available.

## Supplementary Material titles

Table S1. B01A-Antithrombotic agents in the Anatomical therapeutic chemical (ATC) classification

Table S2. ICD-9 and ICD-10 codes of comorbidities

Table S3. Distribution of concomitant medication discontinuation among patients with and without antithrombotic deprescribing and other components of deprescribing

Table S4. Top 20 most frequently used concomitant medications and discontinuation among exposed group

Table S5. Top 20 most frequently used concomitant medications and discontinuation among unexposed group

Table S6. Top 10 frequently used drug combinations and discontinuation among exposed group

Table S7. Top 10 frequently used three-drug combination and discontinuation among unexposed group

---

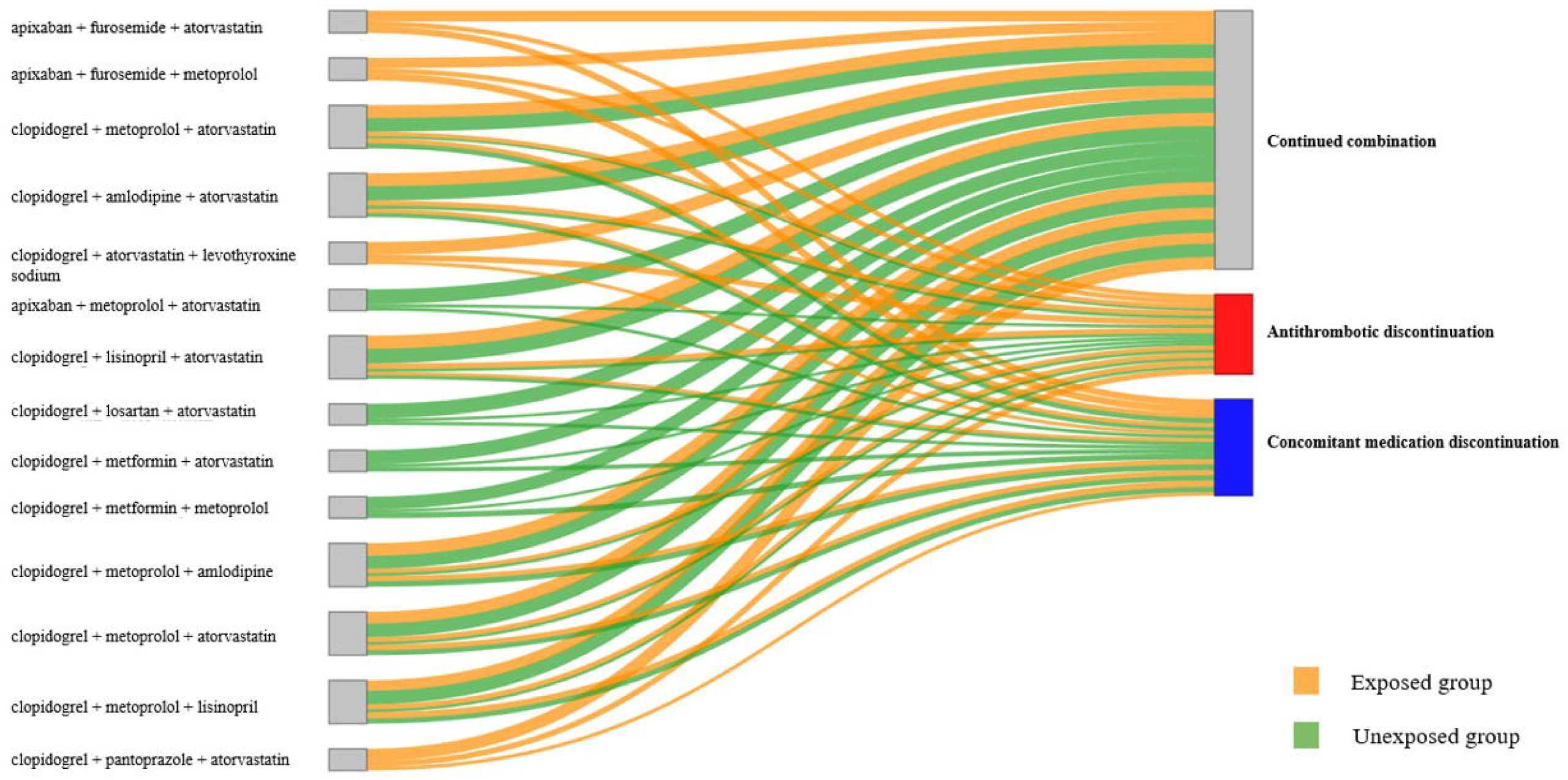

## Notes

### Competing Interest Statement

The authors have declared no competing interest.

### Author Declarations

This study was approved by The Ohio State University Institutional Review Board.

